# Disparities in dolutegravir utilisation in children, adolescents and young adults (0-24 years) living with HIV. An analysis of the IeDEA Paediatric West African cohort

**DOI:** 10.1101/2024.05.24.24307900

**Authors:** Sophie Desmonde, Joycelyn Dame, Karen Malateste, Agatha David, Madeleine Amorissani-Folquet, Sylvie N’Gbeche, Mariam Sylla, Elom Takassi, François Tanoh Eboua, Kouadio Kouakou, Lehila Bagnan Tossa, Caroline Yonaba, Valériane Leroy, IeDEA Pediatric West African cohort

## Abstract

**Introduction:** We describe the 24-month incidence of Dolutegravir (DTG)-containing antiretroviral treatment (ART) initiation since its introduction in 2019 in West Africa.

**Methods:** We included all patients aged 0-24 years on ART from nine clinics in Côte d’Ivoire (n=4), Ghana, Nigeria, Mali, Benin, and Burkina Faso. Baseline varied by clinic and was defined as date of first DTG prescription; patients were followed up until database closure/death/loss to follow-up (LTFU, no visit ≥ 7 months), whichever came first. We computed the cumulative incidence function for DTG initiation; associated factors were explored in a shared frailty model, accounting for clinic heterogeneity.

**Results:** Since 2019, 3,350 patients were included; 47.2% were female;78.9% had been on ART ≥ 12 months. Median baseline age was 12.5 years (Interquartile range[IQR]: 8.4-15.8). Median follow-up was 14 months (IQR: 7-22). The overall cumulative incidence of DTG initiation reached 22.7% (95% Confidence Interval (CI): 21.3-24.2) and 56.4% (95% CI: 54.4-58.4) at 12 and 24 months, respectively. In univariate analyses, those aged <5 years and females were overall less likely to switch. Adjusted on ART line and available viral load (VL) at baseline, females >10 years were less likely to initiate DTG compared to males of the same age (adjusted Hazard Ratio [HR] among 10-14 years: 0.62, 95% CI: 0.54-0.72; among ≥15 years: 0.43, 95% CI: 0.36-0.50), as were those with detectable VL (> 50 copies/mL) compared to those in viral suppression (aHR: 0.86, 95% CI: 0.77-0.97) and those on protease inhibitors compared to those on non-nucleoside reverse-transcriptase inhibitors (aHR after 12 months of roll-out: 0.75, 95% CI: 0.65-0.86).

**Conclusion:** Paediatric DTG uptake was incomplete and unequitable in West African settings: DTG use was least likely in children <5years, females ≥ 10 years and those with detectable viral load. Maintained monitoring and support of treatment practices is required to better ensure universal and equal uptake.

**Key messages:** **What is already known on this topic?**

- Dolutegravir (DTG)-based ART regimens have been recommended as the preferred first-line ART regimens for all individuals living with HIV by the World Health Organisation since 2018, with a specific caution for pregnant women. This was subsequently confirmed for all children with approved DTG dosing and adolescents since 2019.
- The deployment of universal DTG for adults in West Africa has faced challenges including infrastructure challenges, disparities in healthcare system, and initial perinatal safety concerns that significantly impacted women of childbearing age.
- Specific data on DTG uptake in children, adolescents and young adults in West Africa is limited.

**What does this study adds ?**

- This study describes the dynamic of the DTG roll-out over the first 24 months and its correlates since 2019 in a large West African multicentric cohort of children, adolescents and youth.
- We observed a rapid scale-up of DTG among children, adolescents and young adults living with HIV in West Africa, despite the COVID-19 pandemic.
- However, DTG uptake after 24 months was incomplete and inequitable, with adolescent girls and young women being less likely to initiate DTG compared to males, as were those with a detectable viral load (> 50 copies/mL) compared to those in success.
- Younger children < 5 years were also less likely to initiate DTG, explained by the later approval of paediatric formulations and their low availability.

**How may this study affect research, practice or policy?**

- Maintained monitoring, training and updating guidance for healthcare workers is essential to ensure universal uptake to DTG, especially for females, for whom inequity begins at age 10 years.
- Efforts to improve uptake to universal DTG in West Africa require multifaceted interventions including healthcare infrastructure improvement and facilitation of paediatric antiretroviral forecasting and planning.

## INTRODUCTION

Dolutegravir is an integrase strand transfer inhibitor that was first recommended by the World Health Organization (WHO) for antiretroviral therapy (ART) in adults in 2016 [1]. Indeed, DTG-based regimens are highly effective, associated with higher viral suppression rates and higher genetic barrier, reducing potential drug resistance [2]. In 2018, the Tsepamo study in Botswana raised concerns about a potential association between neural tube defects in babies and the use of DTG by mothers at the time of conception [3]. Subsequently, WHO revised recommendations, with a note of caution about using DTG among females of reproductive age [4]. Further data reported a weaker association between DTG and neural tube defects, and modelling studies supported the use of DTG in all people living with HIV because the benefits outweighed the risk [5–7].

In addition, DTG treatment was inaccessible for most children living with HIV, the only formulation available in Sub-Saharan Africa being the 50mg film-coated tablet registered for use in people weighing > 40kg. Data from the Odyssey trial showed that 50 mg DTG tablets given once daily in children provide appropriate pharmacokinetic profiles comparable to adults [8,9]. As a result, the WHO 2019 DTG paediatric dosing guidelines also led to the US FDA approval of 50mg dosing down to 20kg, widening the paediatric population eligible for DTG [4,10,11]. The Odyssey trial team also investigated the dosing of DTG among children receiving rifampicin-containing TB treatment and found twice daily DTG was safe and sufficient, providing a practical ART option for children with HIV-associated TB [12]. Additional data from the trial, addressed the dosing of DTG in children weighing 3kg to <20kg and aged 4 weeks and above, and formulations are now available for children as young as 4 weeks of age [13].

Thus, in 2019, the WHO strongly recommended DTG-containing ART regimens for all adults and adolescents living with HIV, including females of reproductive age regardless of contraception, and in infants and children with approved DTG dosing [14,15].

Since 2019, West African HIV treatment programmes have been transitioning to DTG giving priority to those who need it most (1/those not treated, 2/children receiving non[nucleoside reverse transcriptase inhibitors (NNRTI)-based regimens, 3/children who need to start TB treatment and 4/children receiving LPV/r solid formulations with challenges in administration and achieving viral suppression). To ensure rapid scale-up, WHO recommends DTG switch should occur irrespective of the availability of a viral load test/result, and urge clinicians to ensure sufficient quantification and supply. Of note, paediatric DTG in a 10mg scored formulation to be associated with optimised backbone antiretrovirals (ARVs) such as abacavir/ lamivudine 120/60 mg scored dispersible tablets became available in 2021 in West Africa; paediatric fixed-dose combinations (pALD) were not available during the study period. We sought to describe this early transition, since 2019, among children and adolescents aged 0-24 years in a large multicentric paediatric cohort in West Africa.

## METHODS

### Study design and inclusion criteria

The International epidemiology Database to Evaluate AIDS (IeDEA) paediatric West African Database to evaluate AIDS (pWADA) is aimed at addressing evolving research questions in the field of HIV/AIDS care and treatment using routine data from multicentric HIV/AIDS children cohorts in West Africa [16,17]. This collaboration, initiated in July 2006, involves, in 2024, 10 paediatric referral HIV/AIDS clinics in seven countries: Benin (n=1), Burkina Faso (n=1), Côte d’Ivoire (n=4), Ghana (n=1), Mali (n=1), Nigeria (n=1) and Togo (n=1).

We included all children and adolescents aged 0 – 24 years enrolled in an IeDEA pWADA clinic, with at least one visit from January 2019, time of DTG introduction in West Africa. Those followed up in clinics where DTG was not yet rolled out at the time of the study were excluded from the analysis.

### Outcomes and key definitions

Baseline was site -specific, and defined as the date of the first documented prescription of DTG among patients aged 0-24 years in each site. For those who enrolled after the beginning of DTG roll-out, baseline was date of enrolment. Our main outcome was dolutegravir initiation, which we defined as either transitioning to a DTG-containing regimen from another regimen, or newly initiating DTG-based ART. Start dates for DTG were based on clinician documentation of a new prescription. Competing events were death or loss-to-program. Loss-to-program included patients with a documented transfer to another clinic or those lost-to-follow-up (LTFU), defined as last clinical contact > 7 months at database closeout date. Database closeout dates varied by site, ranging from 2020-2022; site-specific study periods are available in Table 1.

**Table 1.**
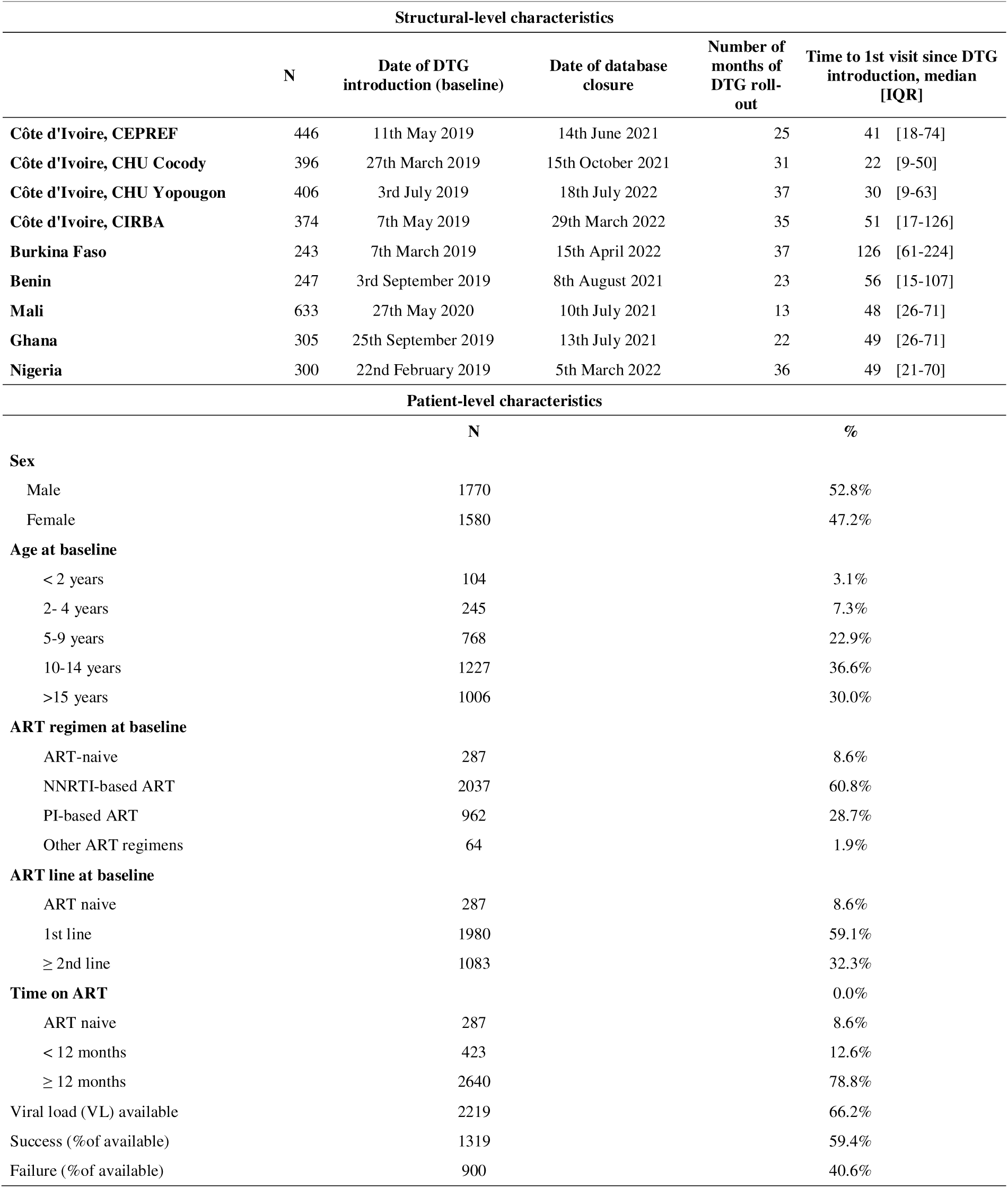
Baseline characteristics of the 9 sites rolling-out DTG and 3,350 patients 0-24 years living with HIV and enrolled in the IeDEA pediatric WestAfrica, 2019-2022.

### Statistical analysis

Patients were followed up from baseline until DTG initiation, a competing event (death, transfer or LTFU), or database closeout date, when they were censored.

First, we calculated crude cumulative incidence proportions over the first 24 months after DTG introduction, along with 95% confidence intervals (CIs), for DTG initiation and competing events using the Aalen-Johansen estimator. Second, we used multi-level survival analysis to estimate hazard ratios for time to DTG initiation [18]. We computed a Cox regression model with mixed effects, incorporating a clinic-specific random effect to account for within-clinic homogeneity in outcomes, and the shared frailty followed a gamma distribution. Model was adjusted for sex stratified by age at baseline, duration on ART, baseline virological status (viral load [VL] unavailable, detectable VL≥ 50 copies/mL or undetectable VL <50 copies/mL) and type of ART regimen immediately prior to DTG initiation according to time.

### Patient and public involvement

This study was conducted using programmatic data collected routinely. Patients were not involved in the analysis plan or result interpretation. Patients did not contribute to the writing or editing of this manuscript.

### Ethics approval

Each participating country formally agreed to contribute paediatric data, with local institutional review board and National Institute of Health approval to contribute to the analyses. Specific individual consent was not required as this was routine collected data.

## RESULTS

Between January 2019 and July 2022, of the 10 paediatric clinical sites contributing to the pWADA cohort, all except the site in Togo had initiated DTG roll-out. Documentation of DTG prescription was first in Nigeria (February 2019) and then in Burkina Faso and the Cocody University Hospital in Cote d’Ivoire (March 2019). Other Ivorian sites accessed DTG between May-July 2019, and the clinical sites in Benin and Ghana began roll-out in September 2019. Lastly, the Gabriel Toure Hospital in Mali began prescribing DTG in May 2020.

Of the 3,891 children with at least one clinical visit since 2019, 3,350 children and adolescents were eligible and included in this study, of whom 3,063 (91.4%) were on ART and in active follow-up at baseline, and 287 (8.6%) ART-naive were newly enrolled during the study period (supplemental content, Figure A). Overall, 47% were female and median age at baseline was 12.5 years (interquartile range [IQR]: 8.4-15.8). Overall baseline characteristics are presented in table 1. Overall, 59.1% were of first-line ART, though this varied by clinic (Supplemental Content, table S1): in Benin, Burkina Faso and the CIRBA in Côte d’Ivoire, >50% were already on second-line regimens, while 80.3% of patients in Ghana were still on first-line. Overall, 78.8% had been on ART > 12 months, median time since ART initiation was 6.9 years (IQR: 3.7-9.9). VL measurement was available at baseline for 66.2% of children, of whom 59.4% were in viral suppression (VL< 50 copies/ml). Viral load measurement at baseline varied across clinics and countries, ranging from 32.9% in Burkina Faso (of whom 47.5% were in viral suppression) to 90.9% at the Yopougon University Hospital, Côte d’Ivoire (of whom 55.0% were in viral suppression).

Overall, since 2019, 1496 (44.7%) patients initiated a DTG-containing regimen, 50 (1.5%) died and 213 (6.4%) were lost to the programme. Median follow-up was 14.0 months [IQR: 7.5-21.9]. Cumulative incidence rates for DTG initiation are described in Figure 1. The 6-month cumulative incidence rate for DTG initiation since roll-out in the clinic was 12.7% (95% Confidence Interval [95%CI]: 11.5-13.9) and reached 56.4% (95%CI: 54.4-58.4) by 24 months. DTG incidence varied greatly according to country (Figure 2). The CIRBA clinic in Côte d’Ivoire was the only site to reach >50%within the first 12 months of DTG introduction (55.7%, 95%CI: 50.5-60.6), followed by Mali (36.5%, 95%CI:32.6-40.4) and the Nigerian Institute for Medical Research (24.1%, 95%CI:19.2-29.2) in Nigeria, while all other clinics were below 20%. After 24 months of roll-out, the CIRBA still had the highest DTG initiation rate (71.4%, 95%CI:66.8-76.0), followed by the Yopougon University Hospital in Côte d’Ivoire (68.1%, 95%CI: 63.4-72.5), the CEPREF clinic in Côte d’Ivoire (65.7%, 95%CI: 59.3-71.4) and the Yaldago Ouedraogo Hospital in Burkina Faso (58.8%, 95%CI: 51.8-65.1), while the other clinics remained below 50%. DTG initiation also varied by sex, with higher cumulative incidence among males compared to females at both 12 and 24 months (Figure 3a). However, we note the gap closing towards the end of the follow-up period.

**Figure 1.**
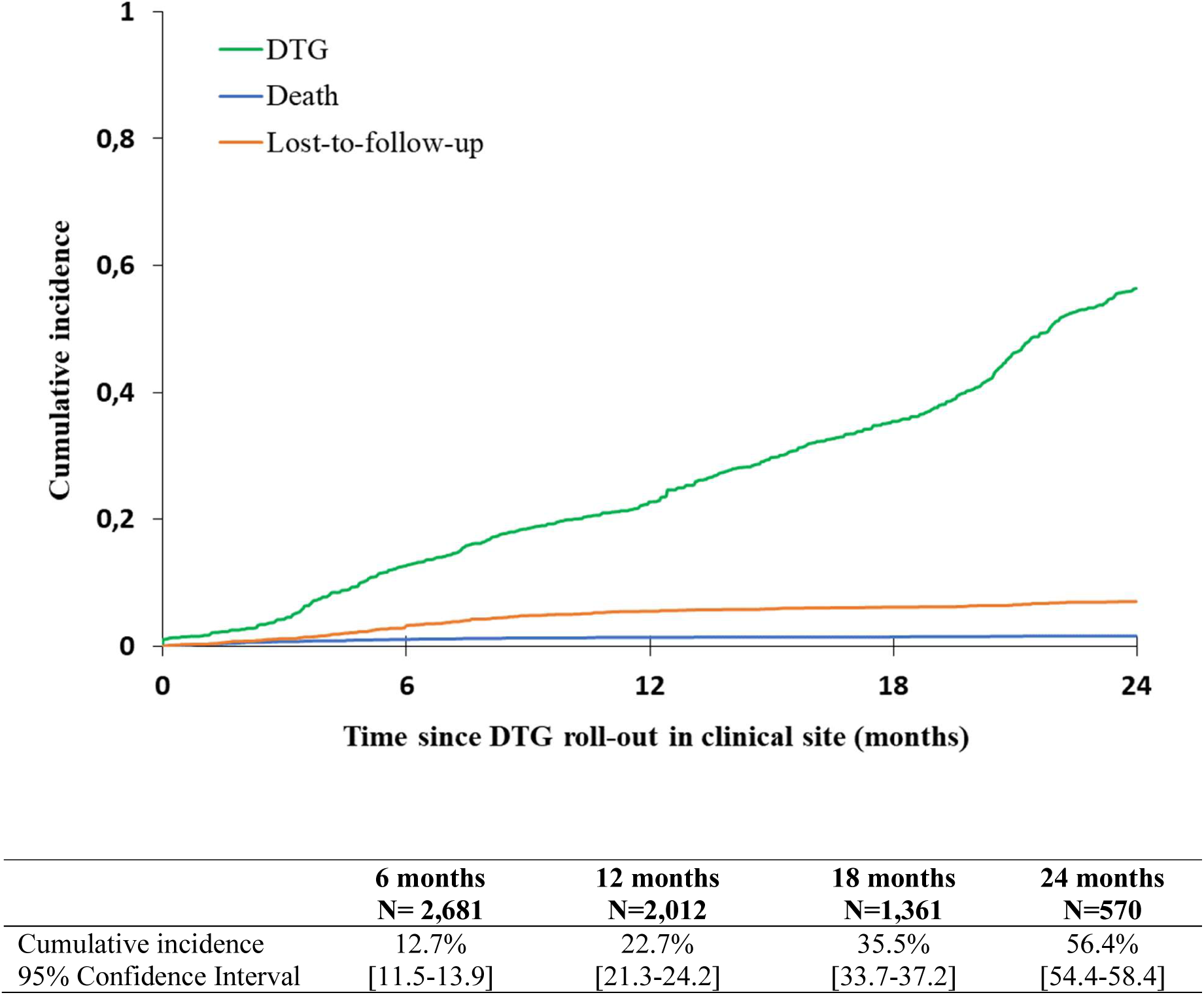
Overall 24-month cumulative incidence function of DTG initiation among the 3,350 children, adolescents and young adults living with HIV and enrolled in the IeDEA pediatric West African sites rolling-out DTG

**Figure 2.**
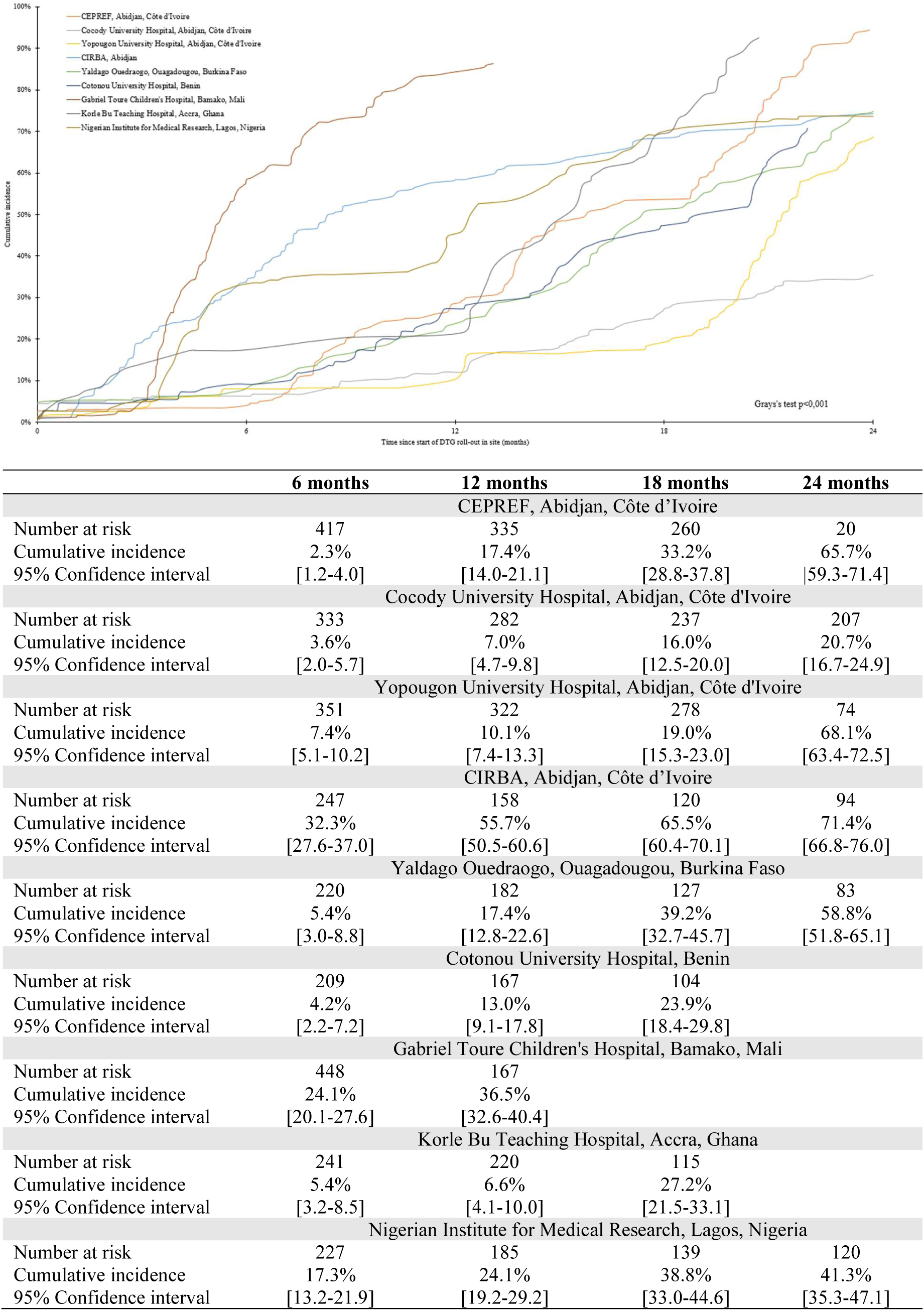
24-month cumulative incidence function of DTG initiation by clinic among the 3,350 children, adolescents and young adults living with HIV and enrolled in the IeDEA pediatric West African clinics rolling-out DTG

**Figure 3.**
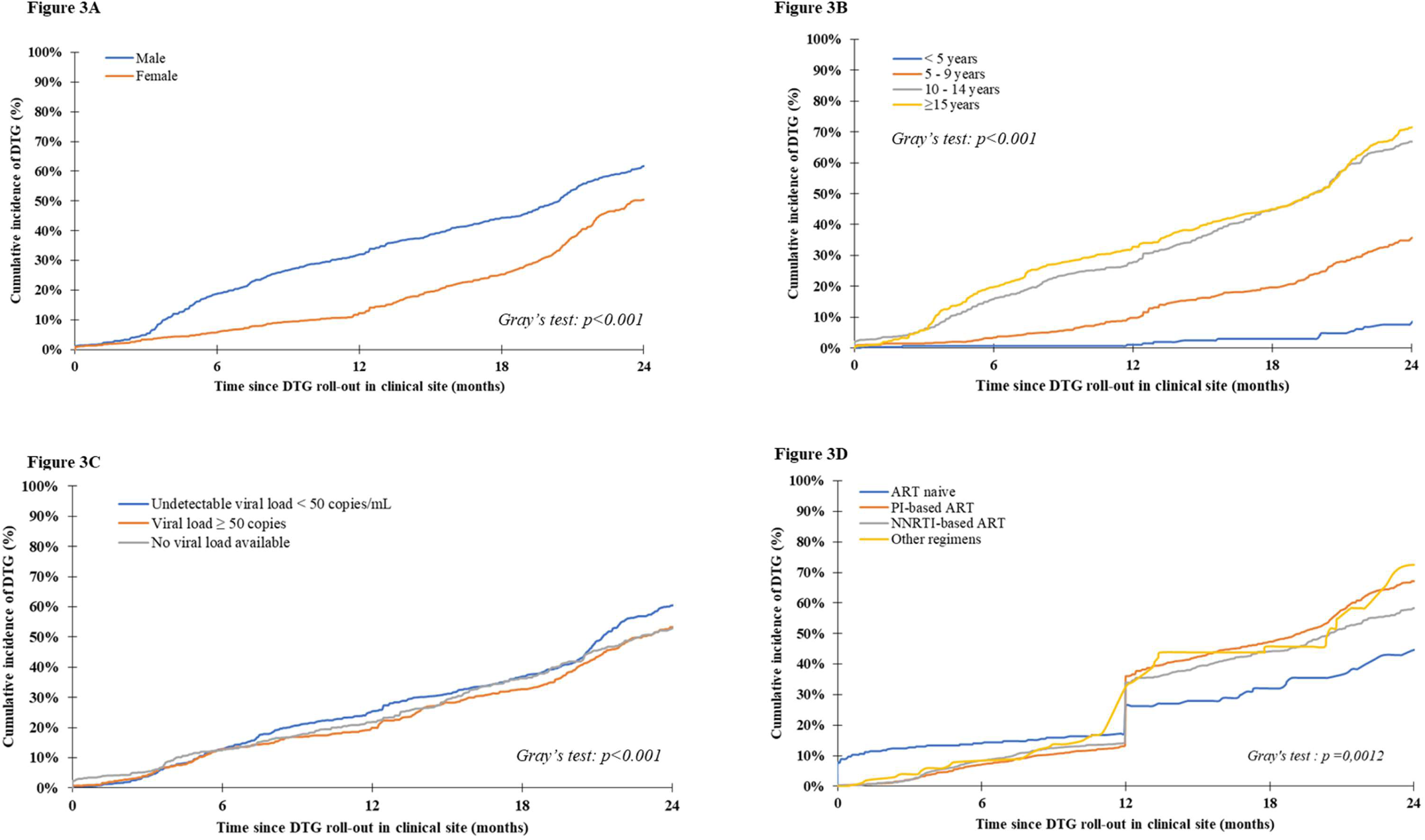
24-month cumulative incidence function of DTG initiation among the 3,350 children, adolescents and young adults living with HIV and enrolled in the IeDEA pediatric West African clinics rolling-out DTG by sex (A), baseline age (B), baseline viral load (C) and baseline ART regimen (D).

Factors associated with DTG initiation are presented in table 2 and Figure 3. In univariate analyses, we found that DTG use was least likely in females, younger children (<5 years), those with detectable viral load and those on PI-based ART. In multivariate analyses, adjusted for all other variables, these associations remained: females were less likely to initiate DTG than their male counterparts. This association strongly depended on age, while there was no difference between sex among those aged <10 years, the adjusted hazard ratio (aHR) was 0.62 [95%CI: 0.54-0.72] among those 10-14 years and 0.43 [95%CI: 0.36-0.50] among those ≥ 15 years. We also found that DTG initiation was less likely among those with detectable viral load (aHR: 0.86, 95%CI: 0.77-0.97) compared to those with viral load < 50 copis/mL. In the first year of DTG roll-out, ART-naïve children were more likely to initiate DTG compared to those on an NNRTI-based ART regimen (aHR:2.00, 95%CI:1.52-2.65); after the first year, those on a PI-based ART regimen were less likely to initiate DTG compared to those on an NNRTI-based ART regimen (aHR: 0.75, 95%CI : 0.65-0.86). We also found that those on ART ≥ 12 months were most likely to switch to DTG compared to those on ART for a shorter duration (aHR:1.29, 95%CI: 1.11-1.51)

**Table 2.**
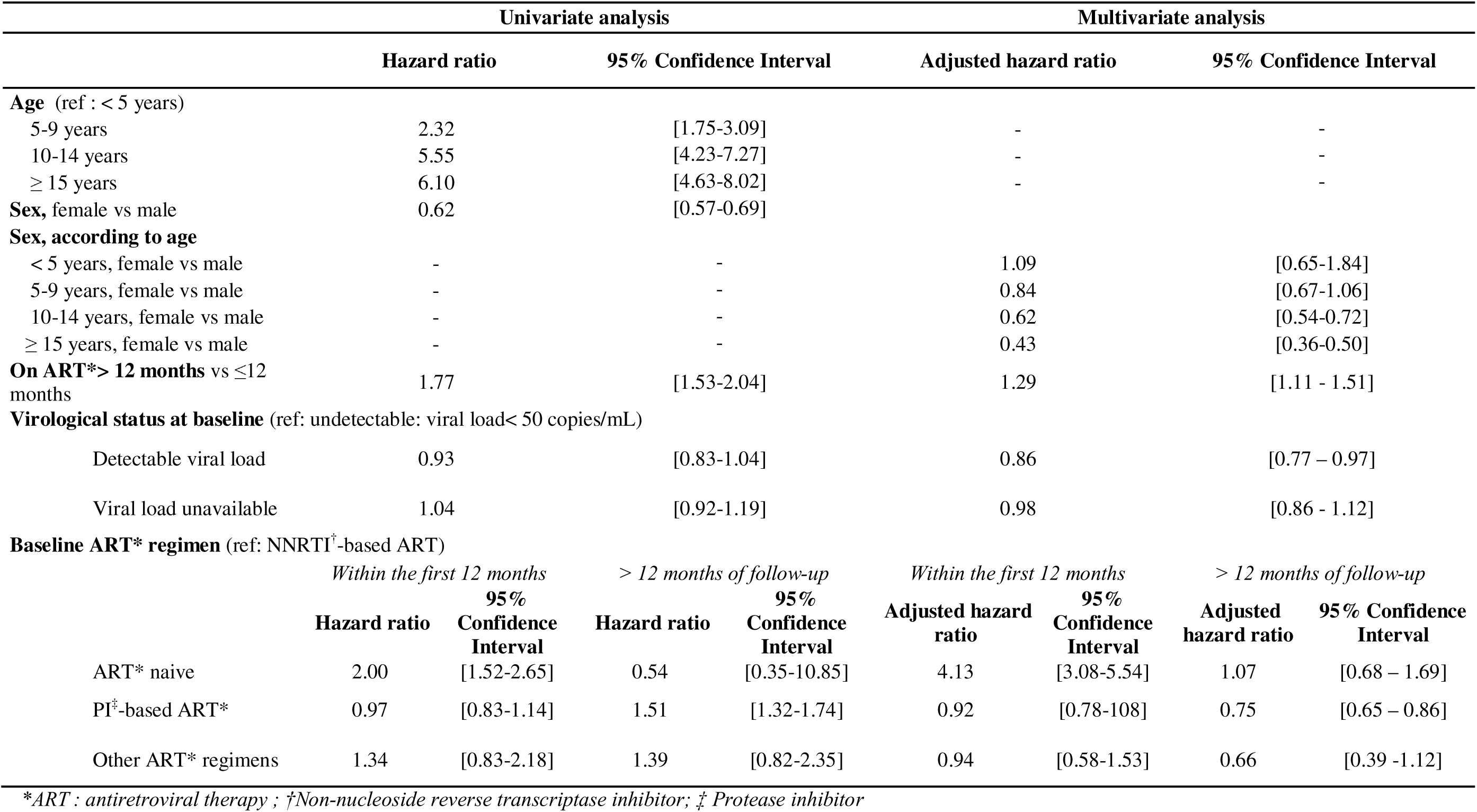
Factors associated with transition to DTG-containing ART regimen among the 3,350 children, adolescents and young adults living with HIV and enrolled in the IeDEA paediatric West African sites rolling out DTG, 2019-2022. Cox regression model with mixed effects, incorporating a clinic-specific random effect and the shared frailty followed a gamma distribution

## DISCUSSION

Universal DTG-containing ART regimens were recommended by the WHO as preferred first-line ART since 2018 in adults and adolescents [4]. This study describes the dynamic of the DTG roll-out over 24 months and its correlates in a large West African cohort of children, adolescents and youth. DTG has been available since 2019 in nearly all clinical sites. Transition to DTG-containing ART regimens was less likely in children <10 years, in females ≥ 10 years and in those having a detectable viral load at baseline. Furthermore, we observed a role of the previous ART regimen at time of DTG roll-out, where use of DTG in the first twelve months was less likely in those on PI-based ART.

First, in our cohort, six out of the seven participating countries had begun prescribing DTG in 2019, among which all but one site (9/10) began in the first semester of 2019. While this is delayed compared to other regions in sub-Saharan Africa, once available[19], we observed a rapid scale-up of DTG roll-out in West Africa, noticeable despite the COVID-19 pandemic in March 2020. Despite the remarkable progress in the roll-out of DTG since 2019, this was also heterogeneous between countries and still incomplete 24 months after the WHO recommendations, as only four clinics reached >50% coverage. As detailed below, population characteristics could explain such differences in transitioning to DTG regimens, but also structural factors not captured in this analysis. We cannot exclude either, an effect of the COVID-19 pandemic that may also have jeopardised and slowed the DTG roll-out in the earlier years [20].

Second, we observed sex disparities among adolescents and young adults ≥ 10 years, where DTG use was less likely in females compared to males of the same age. These disparities increased consistently with age. We assume that clinicians were reluctant in using DTG among female adolescents of childbearing potential because they were aware of the initial signal of neural tube defects reported from Botswana that led to a note of caution in the WHO recommendations in 2018 [3,4]. Since most sites had begun DTG roll-out before the WHO recommended DTG to all, in July 2019, providing further reassurance on DTG due to the declining estimate of neural tube defect risk and observed efficacy, we hypothesise our observation is mostly related to the delay in disseminating such information at the field level [14]. Thus, the observed challenges in DTG use in adolescent and young females also reflects the fear of health care providers to address this concern and their inability to offer contraceptive use in this population. Indeed, contraceptive uptake among adolescent girls and young women in Sub-Saharan Africa is low [21,22] and healthcare providers lack education or face bias in addressing these questions [23,24]. The gap between males and females in terms of DTG initiation tended to close towards the end of the follow-up period. But, this inequity for young females should be addressed urgently and women should be provided with information about benefits and risks to make an informed choice regarding the use of DTG or other ART. Similar studies in the adult population have also reported this trend [25,26]. We advocate that if healthcare providers and stakeholders continue to be educated on the evidence of DTG safety among women of childbearing potential, DTG use in adolescent girls and young women would increase and match that of their male counterparts.

Third, we report that children, adolescents and young adults with detectable viral load were less likely to initiate DTG compared to those in viral suppression in our cohort. This is not in line with the paediatric ARV guidelines recommending that DTG transition should occur irrespective of the availability of viral load test [15]. However, this result is likely driven by the adolescent and young adult population. This is most likely related to anticipated adherence challenges in children and adolescents and inadequate psychological support, which increases the risks of virological failure in this population [27]. Suboptimal adherence to ART has long been identified as a major contributor to the development of drug resistance among people living with HIV [28,29]. Despite the higher genetic barrier in DTG, several studies have reported on the emergence of integrase inhibitor drug resistance and reduced efficacy in patients on DTG receiving functional monotherapy [30,31]. Viral failure in children and adolescents living with perinatally acquired HIV can be either the result of poor adherence, frequent in this population, or also often the consequence of pre-treatment drug resistance caused by suboptimal maternal ART regimens [32]. In the absence of drug resistance testing, clinicians are inclined to prioritise therapeutic education to improve adherence first, despite persistent challenges in this domain, before transitioning to DTG-containing ART regimens. But this can end in a lack of chance too if not switching those who have unknown drug resistance mutations. Affordable technologies for detecting HIV drug resistance among those failing ART would be needed to distinguish between children and adolescents living with HIV with HIV[drug resistance and those who have suboptimal adherence in low-income settings. This would also be important and useful when monitoring patients on DTG, as evidence of pre-treatment resistance to DTG in children and adolescents emerge [33].

Fourth, we report that ART-naive patients were most likely to initiate DTG. This was an expected result since the WHO recommendations in the paediatric population prioritise those not treated [14]. We also found that those on PI-based ART regimens were less likely to initiate DTG compared to those on NNRTI-based ART. Several reasons can explain this expected observation. First, concerns about the development of resistance, as described above, may have led clinicians to favour PI–based regimens if there were anticipated adherence challenges. Indeed, observations from the DAWNING study suggest that failure on PIs is less likely to lead to resistance in adults [34]. Second, countries that had existing stocks of LPV/r pellets or granules were encouraged to consider utilising these stocks, in order to transition to DTG with minimal wastage [35].

Fifth, we found that children, adolescents and young adults on ART ≥ 12 months were most likely to switch compared to those who had just enrolled into care and initiated ART. This observation is particularly interesting in light of the fact that ART-naive clients were also more likely to initiate on DTG, suggesting that DTG is being adopted across both treatment-experienced and naive populations. This dual trend underscores the growing confidence in DTG as a first-line option for ART-naive clients as well as a preferred switch option for patients with longer ART histories.

Finally, we report in univariate analyses that DTG use was least likely children < 5 years. This was an expected result in the early years of our study, since paediatric DTG (pDTG, 10mg) was not widely available then. Cote d’Ivoire, Benin and Nigeria were the first countries to rollout paediatric formulations [36]. Furthermore, the TORPEDO study, carried out in Benin and Nigeria reported high acceptability and preference for pDTG [37]. However, our results report low transition in the population. While this is most probably explained by the fact that these younger children were on LPV/r, remaining in the system, it may also be a result of poor planning and stock-outs. This is of particular concern in terms of outcomes in these younger children living with HIV, who may remain at high risk of virological failure in the context of a slow transition to DTG-containing ART regimens [38]. As paediatric treatment optimisation is focused on the continued scale-up of pDTG, irrespective of viral load, and more recently the introduction of paediatric fixed-dose combination of Abacavir-Lamivudine-Dolutegravir (pALD), it is essential for countries to ensure appropriate forecasting and supply, even for small quantities.

Our study presents several limitations. First, the referral clinics involved in these analyses are not representative of their country, leading to a probably over-estimation of DTG roll-out progress. We also reported no DTG roll-out in Togo, although previous literature has reported on the beginning of DTG roll-out since October 2019 [39]. Second, we report that DTG roll-out began in 2019, shortly before the COVID-19 pandemic, where healthcare provision may have been different from that of standard care, thus affecting structural factors associated with DTG initiation. Third, this study was carried out in the early years of DTG roll-out, with varying database closeout dates ranging 2021-2022. This was also amidst perinatal safety concerns regarding the use of DTG among women of childbearing age and before pDTG was rolled out. We do not account for change in practices over time, however, this less complex model, avoids overfitting and associated limitations in external validity of results. Fourth, the actual reasons from the perspective of healthcare providers for not transitioning patients to DTG-based regimens are not documented. This could help inform strategies to address any barriers and facilitate better access to DTG regimens.

This study however provides useful real-world evidence on the scale-up of DTG in children, adolescents and young adults increasing rapidly since 2019 in West Africa, but still incomplete after 24 months. While we anticipate that sex disparities and the slow transition among the youngest children may fade in the coming years, DTG uptake remains unequal. Lack of viral load and wastage concerns should not be a barrier to DTG initiation. Implementation of DTG regimens should be accompanied with an accelerated scaling up of access to viral load before, and after switch to DTG. Continued monitoring of DTG implementation, its outcomes and better planning of treatment option strategies are required to ensure universal access to all.

## ETHICS

This study was conducted using data from the International epidemiology Databases to Evaluate AIDS (IeDEA) consortium, which collects and manages de-identified patient data from various sites across the globe for research purposes. Ethical approval for the conduct of this study was obtained from the Institutional Review Boards (IRBs) or equivalent ethics committees of each participating country and institution, ensuring compliance with both national and international ethical guidelines for research involving human subjects.

The following IRBs and ethics committees provided approval for the use of data in this study:

- Côte d’Ivoire, Comité National d’Ethique des Sciences de la Vie et de la Sané (CNESVS), IRB00009111, Expiration date: 02/17/2025
- Nigeria, National Inst Med Rsch (NIMR) IRB #1 – Biomedical, IRB00003224, Expiration date: 3/22/2026
- Mali, U Mali Faculty Med Pharmacy & Dentistry IRB #1 – USTTB, IRB00001983, Expiration date: 09/02/2025
- Burkina Faso, Comité d’Ethique pour la Recherche en Santé, IRB000013418, Expiration date: 02/17/2025
- Bénin, Comité National d’Ethique pour la Recherche en Santé (CNPERS), IRB00006860, Expiration date: 6/20/2027
- Togo, Comité de Bioethique pour la Recherche en Santé (CBRS), IRB00009547, Expiration date: 11/03/2026
- Ghana, U Ghana Med Sch/College of Hlth Sciences, IRB00006220, Expiration date: 11/15/2024

All data used were de-identified prior to analysis, ensuring that patient confidentiality and privacy were protected.

## Data Availability

All data produced in the present study are available upon reasonable request to the authors

## CONTRIBUTORSHIP STATEMENT

SD and VL conceived the study, designed the methodology. KM was responsible for data collection and management. SD provided statistical expertise and analysed the data. JD, AD, MAF, SNG, MS, ET, KK, FTE, LBT and CY contributed data and were involved in the interpretation and analysis of the findings, providing input that informed the study’s conclusions and implications. All authors contributed to the drafting of the manuscript, provided revisions, and approved the final version for submission. VL acted as garantor.

## ACKNOWLEDGEMENTS

Research reported in this publication was supported by the US National Institutes of Health (National Institute of Allergy and Infectious Diseases, the Eunice Kennedy Shriver National Institute of Child Health and Human Development, the National Cancer Institute, the National Institute of Mental Health, the National Institute on Drug Abuse, the National Heart, Lung, and Blood Institute, the National Institute on Alcohol Abuse and Alcoholism, the National Institute of Diabetes and Digestive and Kidney Diseases, the Fogarty International Center) under Award Number U01AI069919. The content is solely the responsibility of the authors and does not necessarily represent the official views of the National Institutes of Health

## Site investigators and paediatric cohorts

Paediatric cohorts: Caroline Yonaba, CHU Yalgado Ouadraogo; Lehila Bagnan, CNHU, Cotonou, Benin; Jocelyn Dame, Lorna Renner Korle Bu Hospital, Accra, Ghana; Sylvie Marie N’Gbeche, ACONDA CePReF, Abidjan, Ivory Coast; Kouadio Kouakou, CIRBA, Abidjan, Cote d’Ivoire; Madeleine Amorissani Folquet, CHU de Cocody, Abidjan, Cote d’Ivoire; François Tanoh Eboua, CHU de Yopougon, Abidjan, Cote d’Ivoire; Fatoumata Dicko Traore, Hopital Gabriel Toure, Bamako, Mali; Oliver Ezchechi,, Agatha David, Rosemary Audu, NIMR, Lagos, Nigeria; Elom Takassi, CHU Sylvanus Olympio, Lomé,Togo.

## Regional coordination

Antoine Jaquet (PI), Didier Koumavi Ekouevi (PI), François Dabis, Charlotte Bernard, Caroline Couturier, Karen Malateste, Olivier Marcy, Marie Kerbie Plaisy, Elodie Rabourdin, Thierry Tiendrebeogo: ADERA, University of Bordeaux, National Institute for Health and Medical Research (Inserm) UMR1219, Research Institute for Sustainable Development (IRD) EMR 271, Bordeaux Population Health Centre, Bordeaux, France

Désiré Dahourou, Sophie Desmonde, Gildas Boris Hedible, Julie Jesson, Valeriane Leroy, Emile Sodinyessi: CERPOP, Inserm UMR1295, University of Toulouse, Toulouse, France

Raoul Moh, Jean-Claude Azani, Kadidja Diarra, Jean Jacques Koffi, Maika Bengali, Abdoulaye Cissé, Guy Gnepa, Eric Komena, Séverin Lenaud, Simon Boni, Eulalie Kangah: PAC-CI program, CHU Treichville, Abidjan, Côte d’Ivoire

## Partner institutions

Emory University: Igho Ofotokun (PI), Anandi Sheth, Cecile Delille Lahiri, Chris Martin Washington University: Noëlle Benzekri, Geoffrey Gottlieb, Geneva University: Olivia Keiser

