## Supplemental Table 1 for "Disparities in dolutegravir utilisation in children, adolescents and young adults (0-24 years) living with HIV. An analysis of the IeDEA Paediatric West African cohort"

Table S1 - Patient-level baseline characteristics of the 3,350 patients 0-24 years living with HIV and enrolled in the participating IeDEA pediatric West African clinical , 2019-2022

| Country/Clinics | Côte d'Ivoire CEPREF | | Côte d'Ivoire CHU Cocody | | Côte d'Ivoire CHUYopogon | | Côte d'Ivoire CIRBA | | Burkina Faso CHUYO | | Benin CNHU | | Mali GABRIEL | | Ghana KBTH | | Niegeria NIMR | | *p-value** |
|---|---|---|---|---|---|---|---|---|---|---|---|---|---|---|---|---|---|---|---|
| **Total** | 446 | | 396 | | 406 | | 374 | | 243 | | 247 | | 633 | | 305 | | 300 | | |
| **Date of DTG introduction (baseline)** | 11th May 2019 | | 27th March 2019 | | 3rd July 2019 | | 7th May 2019 | | 7th March 2019 | | 3rd September 2019 | | 27th May 2020 | | 25th September 2019 | | 22nd February 2019 | | |
| **Date of database closure** | 14th June 2021 | | 15th October 2021 | | 18th July 2022 | | 29th March 2022 | | 15th April 2022 | | 8th August 2021 | | 10th July 2021 | | 13th July 2021 | | 5th March 2022 | | |
| **Number of months of DTG roll-out** | 25 | | 31 | | 37 | | 35 | | 37 | | 23 | | 13 | | 22 | | 36 | | |
| **Time to 1st visit since DTG introduction, me** | 41 [18-74] | | 22 [9-50] | | 30 [9-63] | | 51 [17-126] | | 126 [61-224] | | 56 [15-107] | | 48 [26-71] | | 49 [26-71] | | 49 [21-70] | | <.0001 |
| **Sex** | | | | | | | | | | | | | | | | | | | 0,0518 |
| Male | 207 | 46.4% | 209 | 52.8% | 202 | 49.8% | 199 | 53.2% | 124 | 51.0% | 141 | 57.1% | 357 | 56.4% | 170 | 55.7% | 161 | 53.7% | |
| Female | 239 | 53.6% | 187 | 47.2% | 204 | 50.2% | 175 | 46.8% | 119 | 49.0% | 106 | 42.9% | 276 | 43.6% | 135 | 44.3% | 139 | 46.3% | |
| **Age at baseline** | | | | | | | | | | | | | | | | | | | <.0001 |
| Median, [IQR] | 14.3 [9.8-17.3] | | 11.7 [7.3-15.2] | | 14.5 [10.1-17.7] | | 14.3 [11.2-17.6] | | 11.1 [8.1-13.9] | | 12 [7.3-15.2] | | 13.2 [9.1-16.5] | | 9.8 6.0-12.7] | | 10.7 [8.2-12.8] | | |
| < 2 years | 5 | 1.1% | 24 | 6.1% | 7 | 1.7% | 2 | 0.5% | 8 | 3.3% | 12 | 4.9% | 27 | 4.3% | 16 | 5.2% | 3 | 1.0% | |
| 2- 4 years | 23 | 5.2% | 33 | 8.3% | 29 | 7.1% | 20 | 5.3% | 19 | 7.8% | 23 | 9.3% | 49 | 7.7% | 34 | 11.1% | 15 | 5.0% | |
| 5-9 years | 88 | 19.7% | 98 | 24.7% | 63 | 15.5% | 54 | 14.4% | 75 | 30.9% | 57 | 23.1% | 114 | 18.0% | 108 | 35.4% | 111 | 37.0% | |
| 10-14 years | 133 | 29.8% | 138 | 34.8% | 122 | 30.0% | 135 | 36.1% | 97 | 39.9% | 91 | 36.8% | 210 | 33.2% | 134 | 43.9% | 167 | 55.7% | |
| >15 years | 197 | 44.2% | 103 | 26.0% | 185 | 45.6% | 163 | 43.6% | 44 | 18.1% | 64 | 25.9% | 233 | 36.8% | 13 | 4.3% | 4 | 1.3% | |
| **ART regimen at baseline** | | | | | | | | | | | | | | | | | | | <.0001 |
| ART-naive | 31 | 7.0% | 76 | 19.2% | 14 | 3.4% | 1 | 0.3% | 34 | 14.0% | 29 | 11.7% | 43 | 6.8% | 48 | 15.7% | 11 | 3.7% | |
| NNRTI-based ART | 293 | 65.7% | 243 | 61.4% | 266 | 65.5% | 187 | 50.0% | 113 | 46.5% | 95 | 38.5% | 366 | 57.8% | 241 | 79.0% | 233 | 77.7% | |
| PI-based ART | 122 | 27.4% | 77 | 19.4% | 126 | 31.0% | 159 | 42.5% | 93 | 38.3% | 89 | 36.0% | 224 | 35.4% | 16 | 5.2% | 56 | 18.7% | |
| Other ART regimens | 0 | 0.0% | 0 | 0.0% | 0 | 0.0% | 27 | 7.2% | 3 | 1.2% | 34 | 13.8% | 0 | 0.0% | 0 | 0.0% | 0 | 0.0% | |
| **ART line at baseline** | | | | | | | | | | | | | | | | | | | <.0001 |
| ART naive | 31 | 7.0% | 76 | 19.2% | 14 | 3.4% | 1 | 0.3% | 34 | 14.0% | 29 | 11.7% | 43 | 6.8% | 48 | 15.7% | 11 | 3.7% | |
| 1st line | 301 | 67.5% | 242 | 61.1% | 259 | 63.8% | 169 | 45.2% | 109 | 44.9% | 135 | 54.7% | 291 | 46.0% | 245 | 80.3% | 229 | 76.3% | |
| ≥ 2nd line | 114 | 25.6% | 78 | 19.7% | 133 | 32.8% | 204 | 54.5% | 100 | 41.2% | 83 | 33.6% | 299 | 47.2% | 12 | 3.9% | 60 | 20.0% | |
| **Time on ART** | | | | | | | | | | | | | | | | | | | <.0001 |
| ART naive | 31 | 7.0% | 76 | 19.2% | 14 | 3.4% | 1 | 0.3% | 34 | 14.0% | 29 | 11.7% | 43 | 6.8% | 48 | 15.7% | 11 | 3.7% | |
| < 12 months | 95 | 21.3% | 21 | 5.3% | 5 | 1.2% | 89 | 23.8% | 8 | 3.3% | 28 | 11.3% | 170 | 26.9% | 7 | 2.3% | 0 | 0.0% | |
| ≥ 12 months | 320 | 71.7% | 299 | 75.5% | 387 | 95.3% | 284 | 75.9% | 201 | 82.7% | 190 | 76.9% | 420 | 66.4% | 250 | 82.0% | 289 | 96.3% | |
| **Virological status (success : viral load 50 copies).** | | | | | | | | | | | | | | | | | | | <.0001 |
| Viral load (VL) available | 390 | 87.4% | 300 | 75.8% | 369 | 90.9% | 317 | 84.8% | 80 | 32.9% | 92 | 37.2% | 365 | 57.7% | 120 | 39.3% | 186 | 62.0% | |
| Success (%of available) | 240 | 61.5% | 173 | 57.7% | 203 | 55.0% | 208 | 65.6% | 38 | 47.5% | 46 | 50.0% | 216 | 59.2% | 55 | 45.8% | 140 | 75.3% | |
| Failure (%of available) | 150 | 38.5% | 127 | 42.3% | 166 | 45.0% | 109 | 34.4% | 42 | 52.5% | 46 | 50.0% | 149 | 40.8% | 65 | 54.2% | 46 | 24.7% | |
