## Supplementary figures and images for "Disparities in dolutegravir utilisation in children, adolescents and young adults (0-24 years) living with HIV. An analysis of the IeDEA Paediatric West African cohort"

### Supplemental Figure A

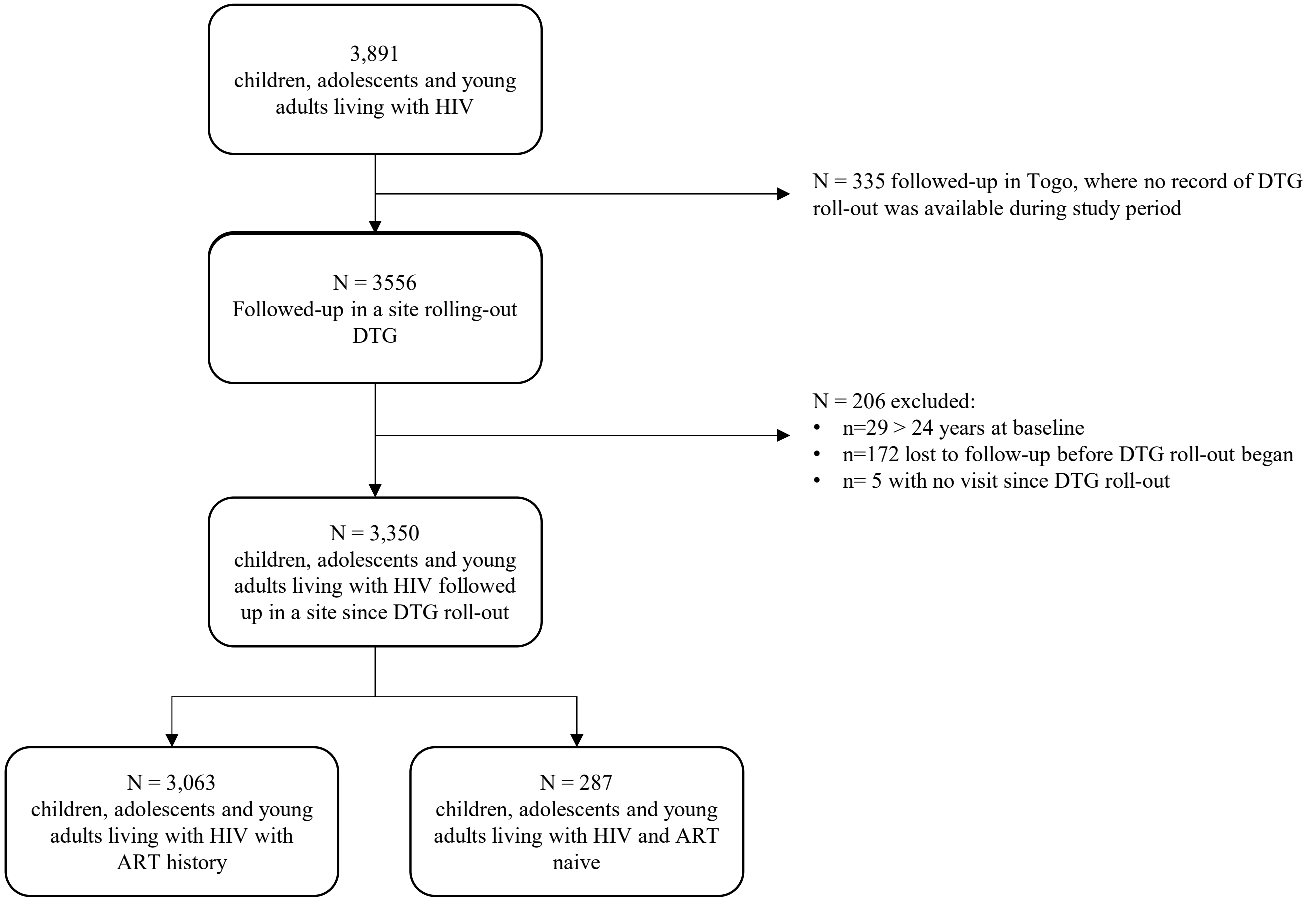


**Figure A** – Patient inclusion flow diagram
